# IL-2 stromal signatures dissect immunotherapy response groups in non-small cell lung cancer (NSCLC)

**DOI:** 10.1101/2021.08.05.21261528

**Authors:** James Monkman, Honesty Kim, Aaron Mayer, Ahmed Mehdi, Nicholas Matigian, Marie Cumberbatch, Milan Bhagat, Rahul Ladwa, Scott N Mueller, Mark N Adams, Ken O’Byrne, Arutha Kulasinghe

## Abstract

**Introduction:** Immunotherapies, such as immune checkpoint inhibitors (ICI) have shown durable benefit in a subset of non-small cell lung cancer (NSCLC) patients. The mechanisms for this are not fully understood, however the composition and activation status of the cellular milieu contained within the tumour microenvironment (TME) is becomingly increasingly recognised as a driving factor in treatment-refractory disease.

**Methods:** Here, we employed multiplex IHC (mIHC), and digital spatial profiling (DSP) to capture the targeted immune proteome and transcriptome of tumour and TME compartments of pre-treatment samples from a 2^nd^ line NSCLC ICI-treated cohort (n=41 patients; n=25 responders, n=16 non-responders).

**Results:** We demonstrate by mIHC that the interaction of CD68^+^ macrophages with PD1^+^, FoxP3^+^ cells is significantly enriched in ICI refractory tumours (p=0.012). Our study revealed that patients sensitive to ICI therapy expressed higher levels of IL2 receptor alpha (CD25, p=0.028) within the tumour compartments, which corresponded with the increased expression of *IL2* mRNA (p=0.001) within their stroma, indicative of key conditions for ICI efficacy prior to treatment. *IL2* mRNA levels within the stroma positively correlated with the expression of pro-apoptotic markers cleaved caspase 9 (p=2e-5) and BAD (p=5.5e-4) and negatively correlated with levels of memory T cells (CD45RO) (p=7e-4). Immuno-inhibitory markers CTLA-4 (p=0.021) and IDO-1 (p=0.023) were also supressed in ICI-responsive patients. Of note, tumour CD44 (p=0.02) was depleted in the response group and corresponded inversely with significantly higher stromal expression of its ligand *SPP1* (osteopontin, p=0.008). Analysis of differentially expressed transcripts indicated the potential inhibition of stromal interferon-gamma (IFNγ) activity, as well as estrogen-receptor and Wnt-1 signalling activity within the tumour cells of ICI responsive patients. Cox survival analysis indicated tumour CD44 expression was associated with poorer prognosis (HR=1.61, p=0.01), consistent with its depletion in ICI sensitive patients. Similarly, stromal CTLA-4 (HR=1.78, p=0.003) and MDSC/M2 macrophage marker ARG1 (HR=2.37, p=0.01) were associated with poorer outcome while levels of apoptotic marker BAD (HR=0.5, p=0.01) appeared protective. Interestingly, stromal mRNA for E-selectin (HR=652, p=0.001), *CCL17* (HR=70, p=0.006) and *MTOR* (HR=1065, p=0.008) were highly associated with poorer outcome, indicating pro-tumourigenic features in the tumour microenvironment that may facilitate ICI resistance.

**Conclusions:** Through multi-modal approaches, we have dissected the characteristics of NSCLC and provide evidence for the role of IL2 and stromal activation by osteopontin in the efficacy of current generations of ICI therapy. The enrichment of SPP1 in the stroma of ICI sensitive patients in our data is a novel finding, indicative of stromal activation that may aid immune cell survival and activity despite no clear association with increased levels of immune infiltrate.

## Introduction

Lung cancer is the leading cause of cancer related mortality in men and second cause of cancer mortality in women, with 5-year overall survival (OS) rates between 10-20% ^1^. Of these cases, long-term benefit to PD-1/PD-L1 immune checkpoint inhibitors (ICIs) are observed in only 20-30%. There is currently an unmet clinical need for predictive biomarkers for ICI therapy ^2–4^. Both membranous tumour cell expression of PD-L1 ^5^ and tumour mutation burden (TMB) ^6, 7^ are FDA approved companion diagnostic biomarker tests used to stratify patients for immunotherapy, however these appear to be independent variables in their predictive capacity ^6, 8, 9^. Difficulties in the implementation and assessment of these assays suggests that much remains to be discovered to understand the biological cues that dictate ICI response.

Both therapy response and tumour progression are governed by tumour intrinsic features and their inherit interaction with the cellular composition of the tumour microenvironment (TME) ^10^. Such interactions activate or supress inflammatory signalling and immune cell recruitment, disrupt antigen presentation, remodel the extracellular matrix (ECM), modify nutrient and oxygen supply and clearance, and overall disrupt the homeostasis that would otherwise allow tumours to be targeted by the host immune system ^4^. Of the milieu of cell subsets within a patient’s TME, very little is understood of their interaction and how these associations may influence both outcome to immunotherapy.

While the field of immunology has benefited from flow cytometry to delineate the roles of cellular hierarchies found in peripheral blood, similarly high-plex methods to analyse both composition and spatial organisation of cells in tissue have been lacking. As such, information garnered thus far has been limited to targeted multiplex immunohistochemistry panels to evaluate prognostic and predictive value of small numbers of cell markers in parallel including CD3^+^, CD4^+^, CD8^+^, FoxP3^+^, PD-1^+^, CD68^+^, PD-L1^+ 11–14^. Interestingly, PD-L1 expression within the immune cell compartment is being increasingly reported and provides confounding evidence to the PD-1/PD-L1 paradigm of tumour-centric immune evasion ^15–18^.

Advances in multiplexed techniques combined with simultaneous imaging readout to measure the composition of tumour and the TME have begun to unravel the cellular phenotypes that associate with therapy response. Here we adopted a discovery approach to examine the cellular composition of lung cancer patient samples using spatial proteomic and transcriptomic methodologies to evaluate tumour and TME compartments independently, with additional single cell level multiplex immunohistochemistry (mIHC). A second line ICI immunotherapy cohort (IO) was examined by targeted 1800 mRNA and 68 protein DSP assays, aswell as 6-plex mIHC. We sought to use unbiased statistical approaches to inform upon on both response to therapy and OS.

## Materials and Methods

This study has Queensland University of Technology (QUT) Human Research Ethics Committee Approval (UHREC #2000000494). Pre-treatment NSCLC tissue microarrays (TMAs) were provided by TriStar Technology Group (USA). The cohort consisted of tumours from patients who received second line ICI immunotherapy (IO cohort). Serial sections of the IO cohort were analysed by Nanostring GeoMX Digital Spatial Profiler (DSP) protein and mRNA panels (Cancer Transcriptome Atlas panel, CTA), and by multiplex immunohistochemistry (mIHC). Clinical endpoints included ICI response status and OS for the IO cohort. All patient clinicopathological, treatment and survival parameters were recorded and provided by TriStar Technology Group.

### Nanostring GeoMX Digital Spatial Profiler (DSP)

NSCLC TMA slides were stained and analysed by DSP at the Systems Biology and Data Science Group at Griffith University (Gold Coast, Australia). Morphology/visualisation markers consisted of CD3, CD68, and Pan-cytokeratin. mRNA panel consisted of 1812 curated genes from the human cancer transcriptome assay (CTA) including housekeeping and negative control probes. Slides were processed and hybridised according to the manufacturer’s instructions. ROI selection on each TMA core was performed such that 660 µm circles were segmented into cytokeratin^+^ (tumour) and cytokeratin^−^ (stroma) regions. Barcodes from these regions were collected to generate measurements per compartment. Barcodes were sequenced, mapped, and counted by NGS readout as per manufacturer’s instructions. Quality control (QC) was performed within the DSP analysis suite to remove outlying probes and collapse counts from 5 probes per gene to single measurements. This QC data was output for bioinformatic analysis.

Protein panel consisted of 68 antibodies including core panels across Human immune cell profiling, IO drug target, Immune activation, Immune cell typing, pan-tumour, cell death, PI3K/AKT panels. Slides were processed as per manufacturer’s instructions and tumour/stroma regions demarcated as above. Antibody barcodes were counted on Ncounter platform as per manufacturer’s instructions and External RNA Controls Consortium (ERCC) QC performed in DSP analysis suite prior to outputting data for bioinformatic analysis.

### Multispectral Multiplex IHC

Slides were stained using validated MOTIF lung cancer PD-1/PD-L1 panel (Akoya Biosciences, US) as per manufacturer instructions on Leica Bond RX (Leica biosystems, US) at the Walter and Eliza Hall Institute (WEHI) histology core (Melbourne, Australia). Whole slide scans were performed on Vectra Polaris (Akoya Biosciences, US) by WEHI histology core, and images were spectrally unmixed in InForm (Akoya Biosciences, US) using MOTIF spectral libraries.

### Image analysis

Image analysis was performed by Enable Medicine (US). Nuclear cell segmentation was performed using DeepCell, followed by segmentation dilation ^19, 20^. Cellular protein expression levels were computed from the mean Opal fluorophore intensity for each biomarker. Cell classification was performed by gating on respective fluorescence intensity. Cells were assigned to classes: CD68^+^, (CD68^+^, PD1^+^), (CD68^+^, PD-L1^+^), CD8^+^, (CD8^+^, PD1^+^), (CD8^+^, FoxP3^+^), (CD8^+^, PD1^+^, FoxP3^+^), (FoxP3^+^, PD1^+^), PanCk^+^, (PanCk^+^, PD-L1^+^). Cell frequency was defined by counting the instances of each cell class and normalizing to the total cell count for each core. Cell interactions were defined by shared edges of the Voronoi tessellation generated from the cell centres ^21, 22^. Cell interaction frequencies were calculated by counting the instances of an interaction and normalizing to the total number of interactions in the core. Neighbourhoods were defined using the k-nearest neighbour algorithm (KNN). For each cell, the cell types of the 10 nearest neighbours were assigned as the features of that cell. These features were then run through an unsupervised KNN algorithm and assigned to 10 clusters. The choice of 10 nearest neighbours and 10 clusters was chosen heuristically as parameters that worked well for a wide variety of datasets ^21, 22^. For cross-sample comparisons, the frequency of each cell type/interaction/neighbourhood was normalized against the highest frequency among the cores analysed. The cross-sample comparisons were used to generate dendrograms of cell frequencies/interactions/neighbourhood. T-tests were performed between response status groups for each of these metrics.

### Data analysis

Data analysis was performed by Queensland Cyber Infrastructure Foundation (QCIF, Qld, Australia). Data quality was evaluated by principal component analysis (PCA) and coefficients of variation (CV) assessed to determine suitability of the RUV-III normalisation method ^23, 24^. Differential analysis was performed within DeSeq2 and Limma packages ^25, 26^ and sparse partial least squares-discriminant analysis (sPLS-DA) was performed within mixOmics package ^27^. The Kaplan-Meier survival analysis (KMSA) and tree structured regression models was conducted within R studio ^28^ using Survival, Party and Partykit package ^29–31^ and plots generated by ggplot2 ^32^. Ingenuity Pathway Analysis (IPA®) was used to evaluate upstream regulators of differentially expressed transcripts (QIAGEN Inc., https://www.qiagenbioinformatics.com/products/ingenuitypathway-analysis).

## Results

### Patient Cohorts

The IO cohort (n=41) consisted of patients who received ICI therapy in the second line setting, of which 39% were classified as responsive (R), while 61% were non-responsive (NR) (Table 1). Anti-PD-1 therapies Nivolumab and Pembrolizumab comprised 94% of treatments, with one patient receiving anti-PD-L1 agent Durvalumab. 94% and 32% patients remained alive at follow up time, for responsive, and non-responsive groups respectively. The cohort contained squamous cell and adenocarcinoma NSCLC histology and was comprised of advanced stage III or IV disease at time of ICI treatment.

**Table 1.** IO Cohort Characteristics

| **IO Cohort** | **Non-responder, N = 25** | **Responder, N = 16** |
| --- | --- | --- |
| **Gender** |  |  |
| **F** | 9 (36%) | 5 (31%) |
| **M** | 16 (64%) | 11 (69%) |
| **Age** | 66 (59, 69) | 58 (58, 63) |
| **ICI Treatment** |  |  |
| **Durvalumab** | 0 (0%) | 1 (6.2%) |
| **Nivolumab** | 22 (88%) | 11 (69%) |
| **Pembrolizumab** | 3 (12%) | 4 (25%) |
| **Current Status** |  |  |
| **Alive** | 8 (32%) | 15 (94%) |
| **Deceased** | 17 (68%) | 1 (6.2%) |
| **Histology** |  |  |
| **Adenocarcinoma** | 12 (48%) | 13 (81%) |
| **Squamous Cell Carcinoma** | 13 (52%) | 3 (19%) |

### Multiplex IHC Spatial analysis

The current PD-L1 IHC companion diagnostic for PD-1/L1 axis ICI therapy remains problematic, both in its clinical assessment and biological meaning ^33^. Here were utilised a robust multispectral, multiplexed IHC (mIHC) assay ^34^ to evaluate the frequency and spatial associations of ICI targets PD-1/PD-L1, cytotoxic T-cells (CD8), T-regs (FoxP3), and macrophages (CD68) (Figure 1 A, D). Normalised cellular counts were assessed for each marker and their phenotypic subsets (e.g. CD8^+^, FoxP3^+^) (Figure 1 B, E, Supp Figure 1 A, B), their direct cell: cell interactions (Supp Figure 1 C, D) as well as nearest neighbour classifications (Figure 1 C, F, Supp Figure 1 E, F) with enrichment of each of these features in ICI response groups evaluated (Figure 1G). Neither tumour or immune cell expression of PD-L1, nor T cell expression of PD-1 was enriched in the responding cohorts, and in fact, the frequency of any phenotype alone did not associate with response in our data. Significantly however, the interactions between CD68^+^ and PD1^+^, FoxP3^+^ cells were enriched in ICI refractory tumours (p=0.012), while a trend existed for the interaction between CD8^+^, PD1^+^, FoxP3^+^ exhausted T cells and PanCk^+^, PD-L1^+^ tumour cells in ICI resistant tumours (p=0.073) (Figure 1G). K-nearest neighbour analysis (Figure 1 C, F) did not indicate enrichment of cellular neighbourhoods described by the markers evaluated in this assay (Figure 1 G, Supp Figure 1G).

**Figure 1.**
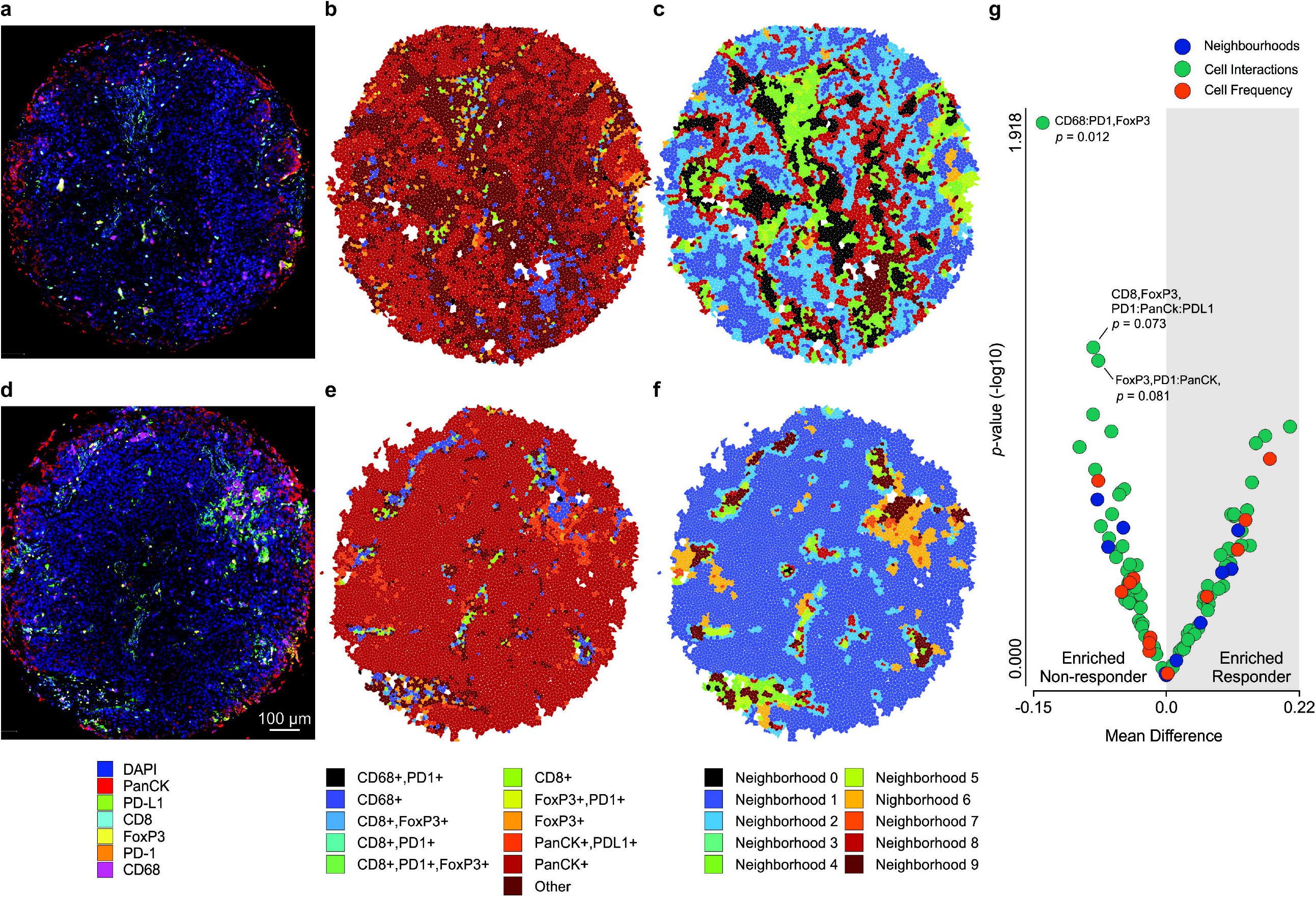
Multispectral spatial analysis revealed the enrichment of cellular interactions in ICI sensitive tumours. Representative analysis of TMA cores from responders (**A-C**) and non-responders (**D-F**). **A** Representative NSCLC core from responder. **B** Concordant cell-type Voronoi. **C** Concordant neighbourhood Voronoi. **D** Representative NSCLC core from non-responder. **E** Concordant cell-type Voronoi. **F** Concordant neighbourhood Voronoi. **G** Volcano plot indicating enrichment and significance of cell frequency, cell interaction and cellular neighbourhoods in responding/non-responding NSCLC cohorts.

### Digital Spatial Profiling

While targeted multiplex IHC offers parallel spatial insights into tumour cellularity, it falls short of the depth required to tease apart the composition of complex tissues. Digital spatial profiling was therefore applied to better delineate the immune proteome and targeted transcriptome of the IO cohort tissues. Regions of tumour cores were segmented using cytokeratin/ non-cytokeratin immunofluorescent masks to enable transcriptome and proteomic marker measurement within tumour and stroma compartments independently (Figure 2). In this way, the measurements obtained from tumour regions comprised both tumour cells as well as infiltrating host cell populations. Similarly, stromal regions described the cellular content of tumour-associated stroma. While most cores contained both tumour and stroma regions, some punches lacked sufficient stromal content for the mRNA assay. As such, of the 41 IO cohort samples, robust transcriptome data was obtained for 33 tumour ROIs and 23 stroma ROIs, which was matched for 21 patients, while protein measurements were complete in both cohorts due to the nature of antibody-based detection compared to that of NGS. The resulting data was assessed for appropriate normalisation, where housekeeping approaches were compared to Remove Unwanted Variation (RUV-III) ^24^ factor analysis which has been demonstrated to be more effective for variance stabilisation than traditional normalisation methods ^35^ (Supp Fig 2). One tumour region of protein assay was excluded from analysis.

**Figure 2.**
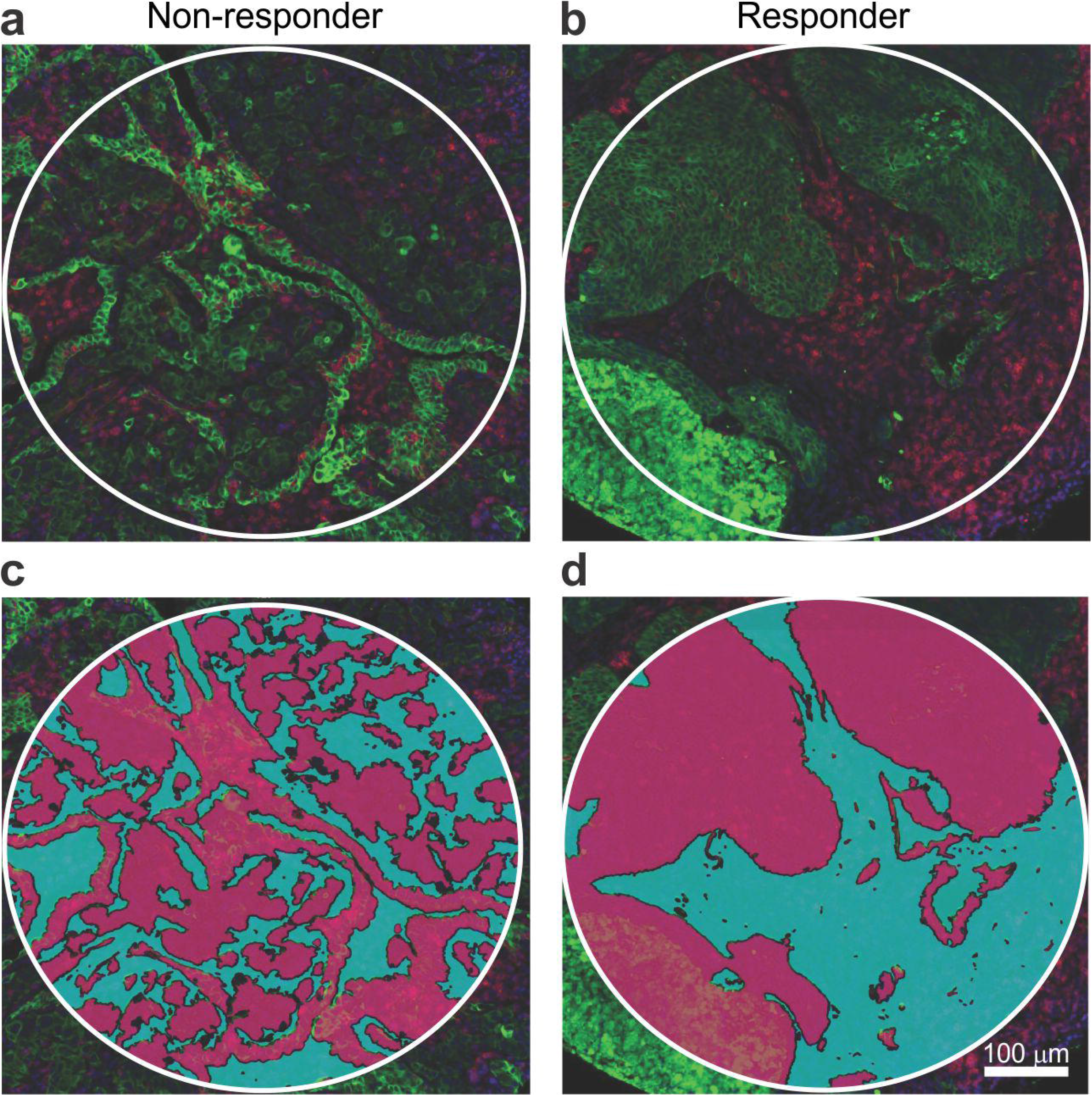
Tumour cores were segmented into PanCK^−^/^+^ regions for DSP analysis of proteomic and mRNA features. **A** Representative field of non-responsive NSCLC tumour. **B** Representative field of responsive NSCLC tumour. **C** Tumour (red) and stroma (aqua) mask applied to tumour shown in A. **D** Tumour (red) and stroma (aqua) mask applied to tumour shown in B. Immunofluorescence markers Green = Pan-cytokeratin, Red = CD45.

Unsupervised dimension reduction was applied to explore the data’s relationship with sample clinical features, including IO response, stage at diagnosis and NSCLC histology, however variance across samples was not clearly attributable to these sample characteristics (Supp Fig 3).

### Postulated markers of IO response by DSP

To further test for associations of cellular markers postulated to be involved in IO response, we evaluated therapeutic targets PD-1 and PD-L1 ^11, 16^, tumour cell antigen presentation (HLA-DR) ^36, 37^ and cytotoxic T cells (CD8^+^) ^12^ in our data. Comparative analysis by DSP alongside our mIHC data indicated that these markers were not significantly associated with ICI response in protein or mRNA DSP assays, despite a trend for higher abundance of PD-1 and CD8 mRNA in tumour regions of ICI sensitive patients (Supp Fig 4).

### Differential analysis of IO response

With this in mind, we sought to identify markers in our DSP data which better discriminated ICI response. Differential analysis was performed between responders (R) and non-responders (NR) for protein markers in tumour regions (R(n)=15, NR(n)=24), stromal regions (R(n)=16, NR(n)=24), as well as mRNA in tumour (R(n)=15, NR(n)=19), and stroma (R(n)=8, NR(n)=18) regions (Figure 3, Supp Table 1).

**Figure 3.**
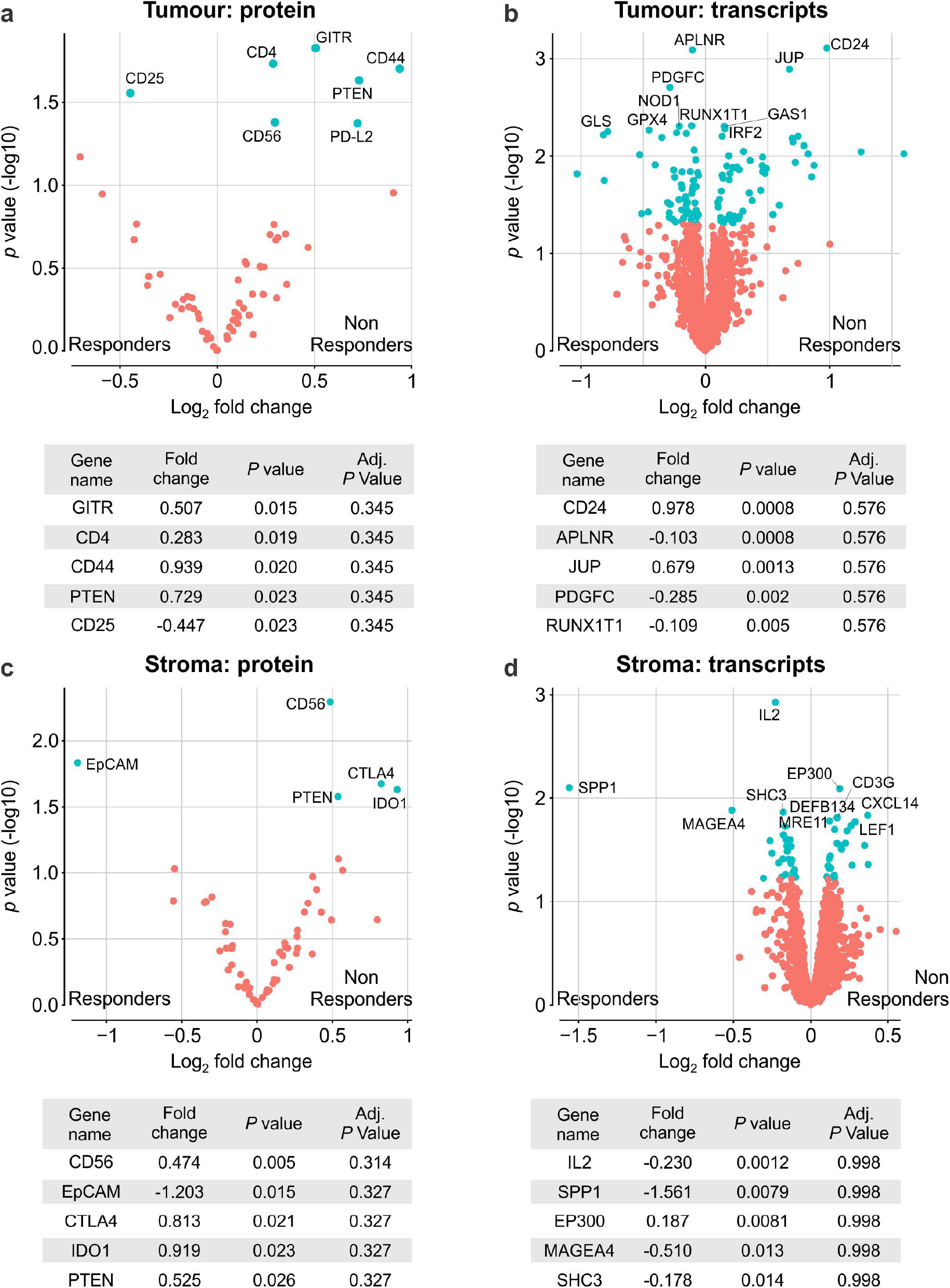
Differential expression of protein and mRNA features by ICI response. **A** Volcano plot of DE proteins in tumour. **B** Volcano plot of DE mRNAs in tumour. **C** Volcano plot of DE proteins in stroma. **D** Volcano plot of DE mRNAs in stroma. Tables indicating most significant DEs shown. Negative value indicates upregulation in responding patients. Differential expression was performed by IO response status within DESeq2 using RUV normalised data.

This analysis indicated the upregulation of IL-2 receptor alpha (CD25) (p=0.028) in tumour regions of responding patients (Figure 3A), which corresponded with higher expression of *IL-2* mRNA (p=0.001) within their stroma (Figure 3D), suggesting key conditions for ICI efficacy. Natural killer cells (CD56) (p=0.005) and immuno-inhibitory markers CTLA4 (p=0.021), IDO1 (p=0.023) were lower in responders’ stroma (Figure 3C), with decreased levels of GITR (p=0.01), CD4 (p=0.01), CD56 (p=0.04), PD-L2 (p=0.04) as well as putative cancer stem cell marker CD44 (p=0.02) and *CD24 mRNA* (p=0.0008) in their tumour regions (Figure 3A, B). Interestingly, the expression of CD44 receptor in tumour regions shared an inverse relationship with the mRNA of its stromal ligand, *SPP1* (osteopontin). Significantly more *SPP1* mRNA (p=0.008) was observed in the stroma of responding patients (Figure 3D), while its tumour cell receptor, CD44, was relatively depleted in these patients. Decreased PTEN was associated with response in both tumour (p=0.02) and stroma (p=0.02). Surprisingly, EPCAM protein (p=0.01) was highly abundant within the stroma of responding patients, and its stromal expression appeared specific as evidenced by a lack of concordant epithelial cell marker, pan-cytokeratin.

IL2 is a key cytokine with pleotropic results on immune cell recruitment, activation, and survival. To investigate the cell phenotypes potentially being influenced by IL2 production, correlations between stromal *IL2* mRNA and concordant stromal protein marker data were evaluated (Supp Fig 5). Interestingly, pro-apoptotic markers cleaved caspase 9 (R=0.73, p=2e-5) and BAD (R=0.63, p=5.5e-4) as well as the IL7 receptor (CD127) (R=0.52, p=7e-3) and IL2 receptor (CD25) (R=0.53, p=5e-3) were positively associated with levels of IL2 mRNA and thus ICI response, while levels of memory T cells (CD45RO) (R=-0.62, p=7e-4) possessed an inverse association.

### ICI Survival Associations

Differentially expressed features were further examined for their prognostic roles given their associations with ICI response (Figure 4, Supp Table 2). Enrichment of tumour CD44 (HR=1.6, p=0.01) and stromal CTLA4 (HR=1.78, p=0.003) and CD56 (HR=1.58, p=0.07) markers in ICI refractory patients corresponded with poorer OS (Figure 4A). Interestingly, non-DE proteins including IL2 mRNA correlate, proapoptotic BAD (HR=0.5, p=0.01), and MDSC/M2 macrophage ARG1 (HR=2.37, p=0.01) markers were associated with improved and poorer outcome, respectively (Supp Table 2). 29 of 109 DE tumour transcripts, as well as 4 of 41 DE stromal transcripts exhibited prognostic associations (Supp Table 2). Segregation of these DE transcripts by their relative expression in ICI responsive patients indicated an inverse prognostic association (Figure 4B, C). Downregulation of several transcripts corresponded with significantly poorer outcome, including stromal E-selectin (SELE) (HR=652, p=8.8e^−4^) and T cell recruitment chemokine CCL17 (HR=70, p=0.006) (Figure 4B). Interestingly, while stromal MTOR expression was not associated with response, it was highly prognostic (HR=1065, p=0.008) (Supp Table 2). Conversely, enrichment of DE transcripts in ICI responsive patients corresponded with significantly enhanced OS, including stromal PLA2G2A (HR=0.03, p=0.02), and tumour NRP1 (HR=0.1, p=0.002), and NOD1 (HR=6e^−4^, p=0.003) (Figure 4B). Taken together, these results indicate the association of these biomarkers with both ICI response and OS outcome following treatment.

**Figure 4.**
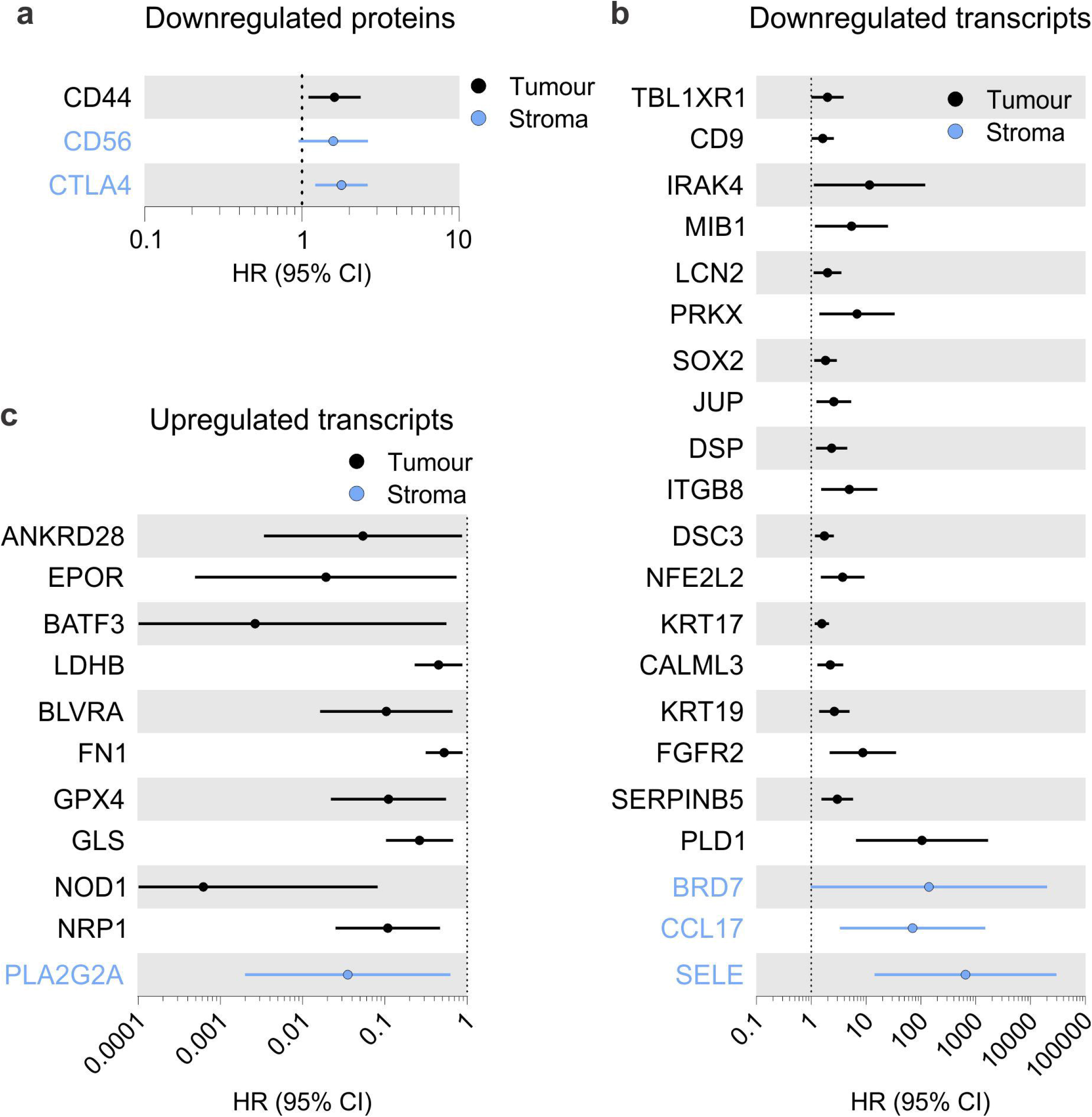
Cox proportional hazard survival analysis of differentially expressed protein and mRNA features by ICI response. **A** Forest plot showing HR and 95% CI of downregulated proteins. **B** Forest plot showing HR and 95% CI of downregulated transcripts. **C** Forest plot showing HR and 95% CI of upregulated transcripts

### Multivariate modelling of IO response

While differential analysis was key in resolving distinguishing features of our data relative to ICI response, their integration may yield an improved diagnostic signature. Multivariate modelling by sparse partial least squares-discriminant analysis (sPLS-DA) was thus performed to discover features that collectively discriminated patient response. sPLS-DA offers a method of feature selection to identify the most predictive or discriminative factors in a dataset that classify samples in a supervised framework, i.e. samples labelled by ICI response ^27^. Features within protein (Figure 5) and mRNA (Figure 6) data were able to efficiently stratify ICI response within tumour and stroma compartments as demonstrated by patient sample separation on components 1 and 2 of the sPLS-DA model (Figure 5 & 6 A-B).

**Figure 5.**
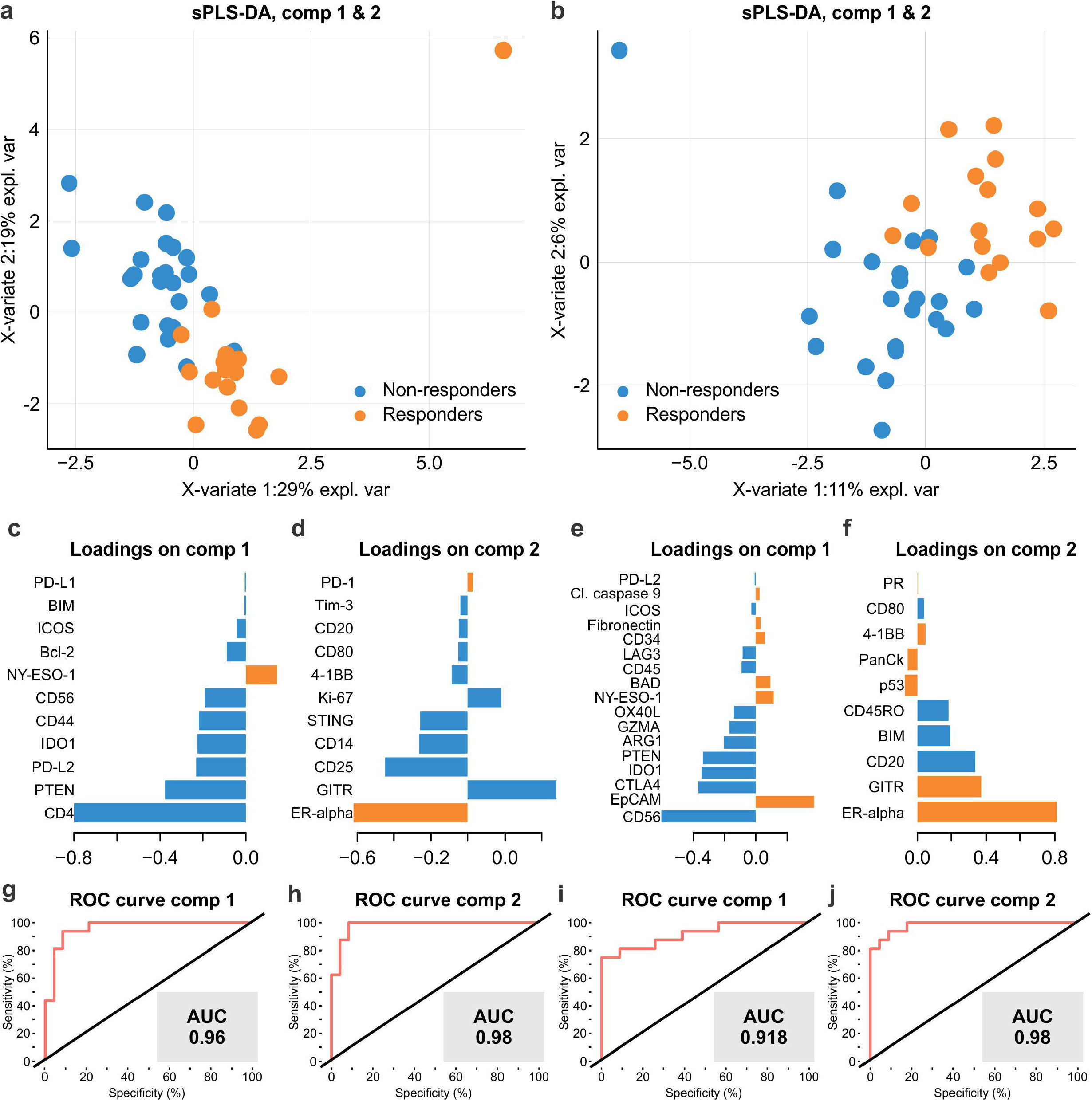
Multivariate sPLS-DA of protein features that distinguish NSCLC IO response. Patient sample discrimination defined by components 1 and 2 of sPLS-DA model for tumour (**A**) and stroma (**B**). **C** Component 1 loading for tumour regions. **D** Component 2 loading for tumour regions. **E** Component 1 loading for stroma. **F** Component 2 loading for stroma. **G** ROC curve to evaluate component 1 tumour signature. **H** ROC curve to evaluate component 2 tumour signature. **I** ROC curve to evaluate component 1 stroma signature. **J** ROC curve to evaluate component 2 stroma signature. **Colour of component loadings indicates patient group in which feature was maximally expressed. Positive or negative values in bar chart indicate positive or negative loading to the discriminant signature**. Blue = Non-responder, Orange = Responder.

**Figure 6.**
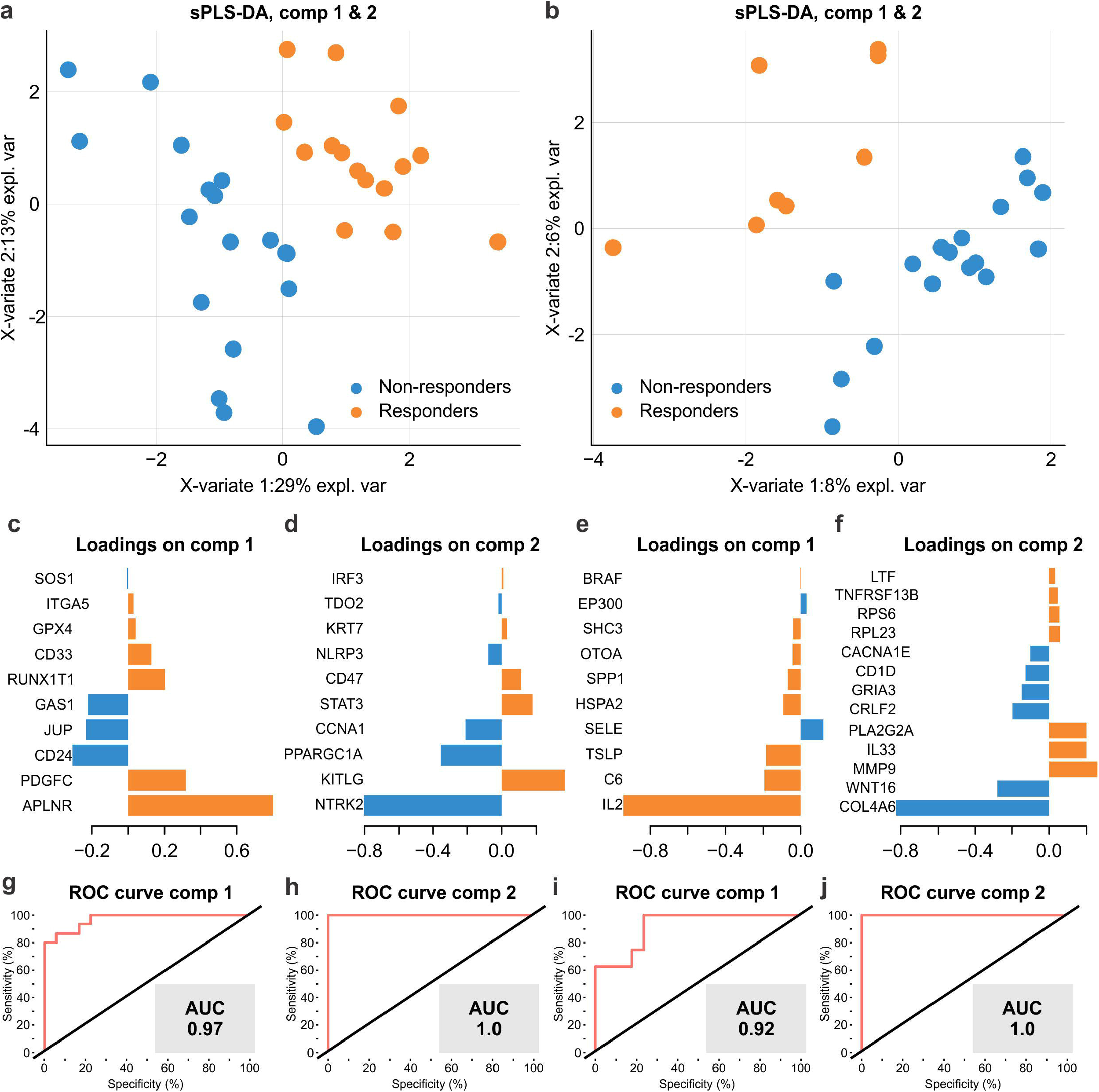
Multivariate sPLS-DA of mRNA features that distinguish NSCLC IO response. Patient sample discrimination defined by components 1 and 2 of sPLS-DA model for tumour (**A**) and stroma (**B**). **C** Component 1 loading for tumour regions. **D** Component 2 loading for tumour regions. **E** Component 1 loading for stroma. **F** Component 2 loading for stroma. **G** ROC curve to evaluate component 1 tumour signature. **H** ROC curve to evaluate component 2 tumour signature. **I** ROC curve to evaluate component 1 stroma signature. **J** ROC curve to evaluate component 2 stroma signature. **Colour of component loadings indicates patient group in which feature was maximally expressed. Positive or negative values in bar chart indicate positive or negative contribution to the discriminant signature**. Blue = Non-responder, Orange = Responder.

Within tumour regions, responding patients could be characterised by increased NY-ESO-1 and lower levels of CD4, PTEN, PDL2, IDO1, CD44 and CD56 markers (AUC=0.96) (Figure 5C) with a secondary signature comprised of higher ER-alpha and lower GITR, CD25, CD14, STING and Ki-67 (AUC=0.98) (Figure 5D). Furthermore, mRNA of responding tumours were characterised by higher *APLNR, PDGFC, RUNX1T1, CD33, GPX4*, and lower levels of *CD24, JUP, GAS1* (AUC=0.97) (Figure 6C). A secondary tumour mRNA signature comprised higher levels of *KITLG, STAT3, CD47* and reduced *NTRK2, PPARGC1A, CCNA1* (AUC=1) (Figure 6D).

Stromal protein markers that indicated IO response comprised higher levels of EpCAM, BAD, NY-ESO-1 and reduced CD56, CTLA4, IDO1, PTEN, ARG1, GZMA and OX40L (AUC= 0.91) (Figure 5E), with a secondary signature composed of increased ER-alpha, GITR, and decreased CD20, BIM, CD45RO (AUC= 0.981) (Figure 5F). Interestingly, *IL2* RNA alone almost solely formed component 1 of the stromal signature (AUC= 0.92) (Figure 6E), while increased *MMP9, PLA2G2A, IL33* and reduced *COL4A6, WNT16, CRLF2, GRIA3*, composed the component 2 signature (AUC=1) (Figure 6F).

Indications from the sPLS-DA together with results of the differential analysis provide evidence for a role of stromal IL2 and its cognate tumour receptor in the efficacy of PD1/L1 ICI therapy. Additionally, lower levels of natural killer cells (CD56) and IDO1 in both tumour and stroma were associated with response. Interestingly, the sPLS-DA model identified that tumour *NTRK2* and stromal *COL4A6* (collagen type IV) expression within the ICI resistant group contributed significantly to the discriminant signature, indicating their potential importance in refractory disease.

### Network and cellular Inference

Ingenuity Pathway Analysis© (IPA®) was used to infer the molecular pathways associated with differentially expressed genes (DEGs) in responding patients. Stromal DEG’s indicated the potential suppression of IFNγ activity (activation score −2.22, p=0.0254) in responders, evidenced by the downregulation of several of its downstream transcriptional products in our data including *CASP3, CCL17, DDX58, HLA-DQB1, IFITM1, PROM1, SELE*, and *VCAM1* (Figure 7A). Tumour DEGs also implicated the inhibition of estrogen receptor (ER) (activation score −2.891, p=0.013) and Wnt-1 (CCN5) (activation score −2.45, p=0.003) signalling pathways in responding patients (Figure 7 B-C).

**Figure 7.**
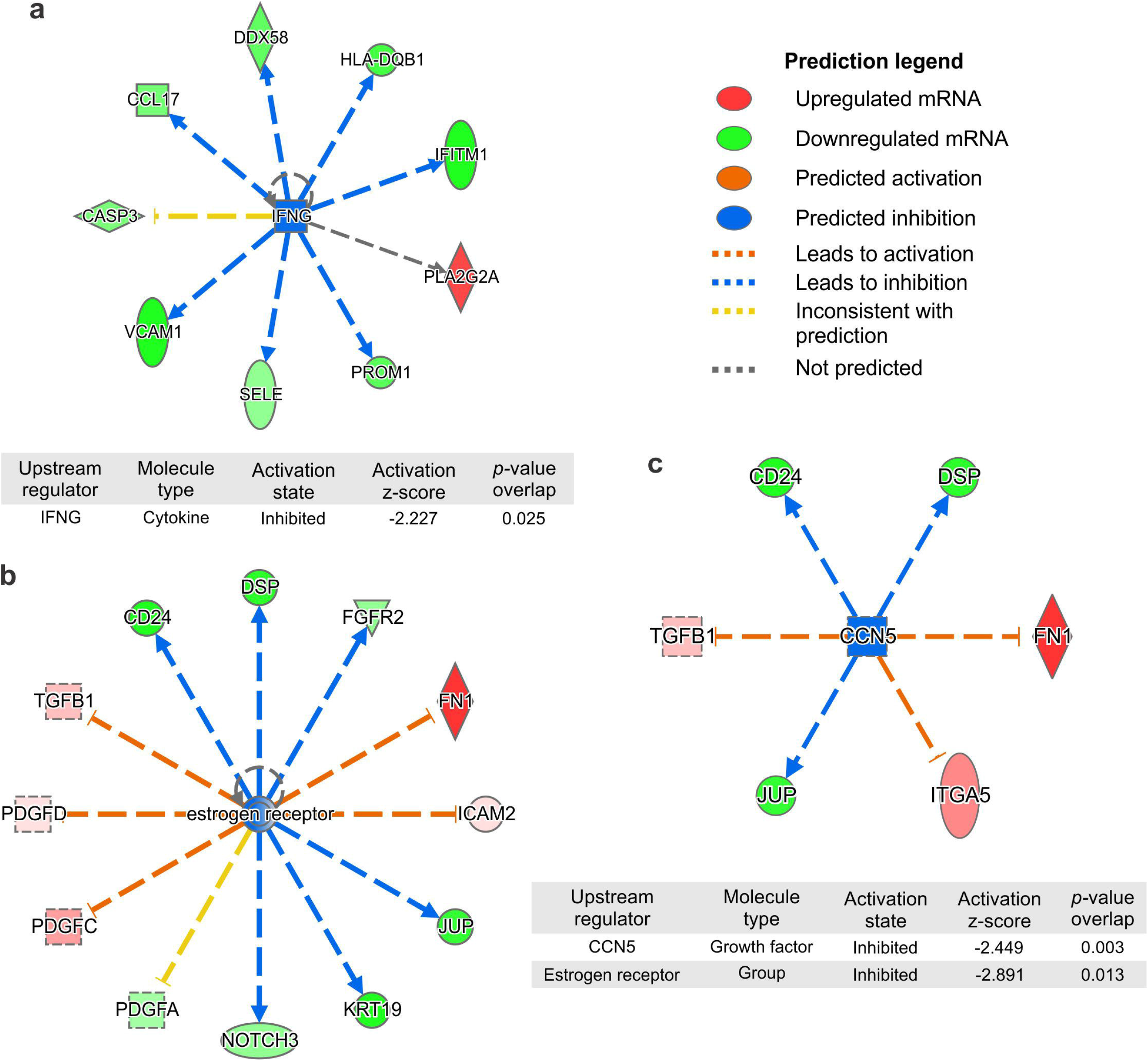
IPA® assessment of differentially expressed genes to infer upstream regulatory pathways. **A** IFNγ pathway activity was predicted to be inhibited within the stroma of responding patients. **B** Estrogen receptor signalling activity was predicted to be inhibited within tumours of responding patients. **C** CCN5 (Wnt-1) signalling was predicted to be inhibited within the tumours of responding patients. **D** Table of signalling pathways predicted to be influenced in responding patients, showing activation z-score and p-value of downstream mRNA target representation. Blue = inhibition, orange = activation, Red=mRNA upregulated, Green = mRNA downregulated

DEG enrichment within the Nanostring CTA pathway annotations showed some associations with the dysregulation of PI3K/Akt signalling, Differentiation, MAPK signalling, Cell Adhesion & Motility, Oxidative Stress, Interleukin signalling, and GPCR signalling in tumour regions (Supp Table 3). Similarly, stromal DEGs were represented in chemokine, GPCR and MAPK signalling pathways (Supp Table 3).

*SPP1* mRNA (osteopontin) was the most highly enriched transcript (Log_2_ FC=1.5) in ICI responsive patients. We therefore sought to assign the cellular source of this transcript in published NSCLC single cell RNAseq data that described nine distinct cell lineages in NSCLC tissue, including the immune cell subsets (T, NK, B, myeloid, MAST) in addition to epithelial cells, fibroblasts, endothelial cells (GSE131907) ^38^. We observed overrepresentation of *SPP1* within the myeloid cell lineage (cluster 1), with more specific expression within monocyte derived macrophages (cluster 5) (Supp Figure 6). The enrichment of *SPP1* mRNA comes despite the absence of increased parental myeloid and macrophage cell markers in our data, including CD11c, CD163, CD68 (Supp Table 1), suggesting myeloid activation and secretion of soluble osteopontin is more directive in ICI response than cellular infiltration alone.

## Discussion

The composition of NSCLC tumours and the interactions that occur at their stromal interface to influence disease progression and ICI response are unclear. Current therapeutic paradigms to reinvigorate cytotoxic anti-tumour responses are hampered by both T cell exhaustion and additional immunosuppressive signals within the TME that predispose a patient’s resistance to therapy. Our study is among the first to characterise these phenomena in both tumour and stromal tissue compartments and provide simultaneous high-plex insight into the in-situ biology of NSCLC patients that subsequently received ICI.

In this study, we sought to profile NSCLC tumours using spatial transcriptomics and multispectral imaging to gain an understanding of the underlying tissue architecture. Overall, the combination of our findings from protein and mRNA differential expression illustrates the landscape of the TME to be consistent with our current understanding of the immune response with several novel findings.

Analysis of stroma indicated that CD56, CTLA4 and IDO1 were significantly associated with resistance to ICI therapy. IDO1 has been shown to be a driver of tumour progression in NSCLC ^39^. It acts to deplete tryptophan, which is required for CD8 T cell proliferation and activation, and acts to reduce CD4 T helper survival ^40^, and is thus an attractive target to sensitise patients to ICI. CTLA-4 is a master inhibitor of T cell activation responsible for regulating self-tolerance, a process which is co-opted in immune evasion ^41^ and was thus the target of first generation ICIs which displayed higher adverse events relative to current PD-1 blockade ^42^. Interestingly EPCAM was also enriched in stroma of responding patients, and is a novel finding that requires further validation, and provides a confounding observation to that made for tumour adjacent stromal EPCAM expression and its association with more aggressive prostate cancer ^43^.

Lower tumour abundance of protein markers GITR, CD4, CD44, CD56, and PD-L2 but not PD-L1 was significantly associated with response, while responders demonstrated increased IL2 receptor alpha (CD25) with concordant expression of the mRNA for its cytokine ligand within the stroma. Of note, limited other mRNAs were upregulated in responders but included *SPP1* which was highly enriched in stroma of responding patients, along with small increases in SHC3 and MAGEA4.

The relative enrichment of cytokine *SPP1* (osteopontin) mRNA within stroma of responding patients (Log_2_ FC 1.5) in our data is novel and suggests that it may have an important role in generating an ICI sensitive niche in tumours. Osteopontin is predominantly expressed and secreted by CD11b^+^ myeloid cells and has been implicated as an immune checkpoint that inactivates cytotoxic activity of CD8 T cells through interaction with the immune CD44 receptor ^44^. Through publicly available scRNAseq data, we show that *SPP1* is highly enriched within monocyte derived macrophage populations and may indicate the specific secretion of this cytokine by activated macrophages. Within LUAD TCGA data, *SPP1* expression was higher in EGFR mutant NSCLC tumours, and was associated with poorer prognosis, with GSEA indicating immunosuppression in high *SPP1* expression group with lower CD8 infiltration and higher M2 macrophage infiltration ^45^. It has been shown to interact with both beta integrins and CD44 on proinflammatory T helper cells. This drives IL-17 production while supressing IL-10, in addition to inducing the hypomethylation of *IFN*γ, promoting its production by T cells, thereby enhancing immune cell survival ^46^. Its upregulation in our data suggests its role is more closely aligned with the latter function and highlights the dichotomous roles that cytokines may have in regulating immune cell activation and survival depending on cellular context.

Interleukin-2 is a potent cytokine that has pleiotropic effects on the immune system including Treg maintenance, T cell differentiation, CD8 T-cell expansion and cytotoxic activity of CD8 and NK cells. Its small but significant upregulation within tumour associated stroma in our data may indicate fertile ground for immune cell survival and tumour clearance. It is produced largely by antigen stimulated CD4^+^ cells, while also produced by CD8, NK and activated DCs. Synergistic activity of IL2 with PD-1 checkpoint inhibition has been demonstrated to enhance CD8 expansion and clearance of pre-clinical viral infection models ^47^ by inducing CD127^+^ and CD44^+^ memory T cell phenotype despite concomitant increase in Tregs. Low dose IL2 has been successfully used to enhance T cell responses in treatment of renal cell carcinoma and metastatic melanoma ^48, 49^ however is subject to variable patient tolerance. More recent work to develop a masked IL2 cytokine that is proteolytically cleaved and activated by tumour-associated MMPs to promote CD8 cell expansion and overcome resistance to ICI has been demonstrated ^50^. Similarly, a tumour targeting recombinant antibody linked to IL2 that localises to tumour cells has been shown to induce cytotoxic CD8 expansion and overcome resistance to ICI ^51^. Thus, we provide here exciting supporting evidence for the therapeutic value of IL2 within the tumour microenvironment and its requirement in the efficacy of PD-1/L1 ICI therapy.

The indication that IFNγ activity is supressed in the tumour-associated stroma of patients that respond to ICI following tumour resection provides a novel, if confounding, insight for the role of this pleiotropic cytokine. IFNγ is typically produced by immune cell subsets including T-cell, NK, Tregs and B cells, however its distinct activities are modulated by the profile of co-secreted soluble cytokines, including IL2. IFNγ has been shown to supress Treg activity to allow expansion of T cells and promote PD-1 response in mouse models ^52^. Conversely, IFNγ has also been shown to prevent CD8 expansion ^53^ and promote T cell apoptosis ^54^. Exhausted CD8 T cells lose secretion of IL2 and IFNγ and feature low proliferation and high expression of inhibitory markers including PD1, LAG3, TIM-3 ^55^. Thus, ICI therapies targeting PD-1 typically induce IFNγ through reinvigoration of T cells and activation NK cells. Recent discovery of a distinct dysregulated CD8^+^ TIL population in the TME that demonstrated higher levels of activation, proliferation and apoptosis with decreased IFNγ production appeared to expand through tumourigenesis and associated with advanced clinical stage and promoted ICI resistance in both clinical and pre-clinical samples ^56^.

In addition, through spatial analysis of multiplex IHC, we show the association of the interactions between macrophages and PD1, FoxP3 cells to be significantly enriched in ICI refractory tumours. These interactions between immune populations are indicative of the paracrine signalling that occurs to drive macroscopic phenotypes and suggests the importance of the next generation of biomarkers that overcome reliance on cell abundance alone.

Overall, our study forms a unique insight into the properties of NSCLC tumours prior to ICI therapy, and discerns features that distinguish subsequent patient response. The ability to measure both immune proteome and transcriptome of FFPE tumour tissue routinely taken during biopsy or resection provides an opportunity to study tumour cellularity at unprecedented depth. We show here in a pilot cohort the strength of such applications in delineating the cues responsible for patient outcome. We identify that while cellular infiltration alone in TMA cores does not associate with ICI response, several key conditions may be required for a robust immune activation upon ICI treatment. While further interrogation and validation of several findings here are required to make conclusive observations, our study forms a basis for the application of next generation technologies for the next generation of diagnostic pathologies.

## Data Availability

The datasets generated during and/or analysed during the current study are available from the corresponding author on reasonable request.

## Acknowledgements

The authors acknowledge the researchers in the Systems Biology and Data Science Research Facility (Griffith University), the Walter and Eliza Hall Institute (WEHI) histology core and Enable Medicine.

## Supplementary Figures

**Supp Figure 1.**
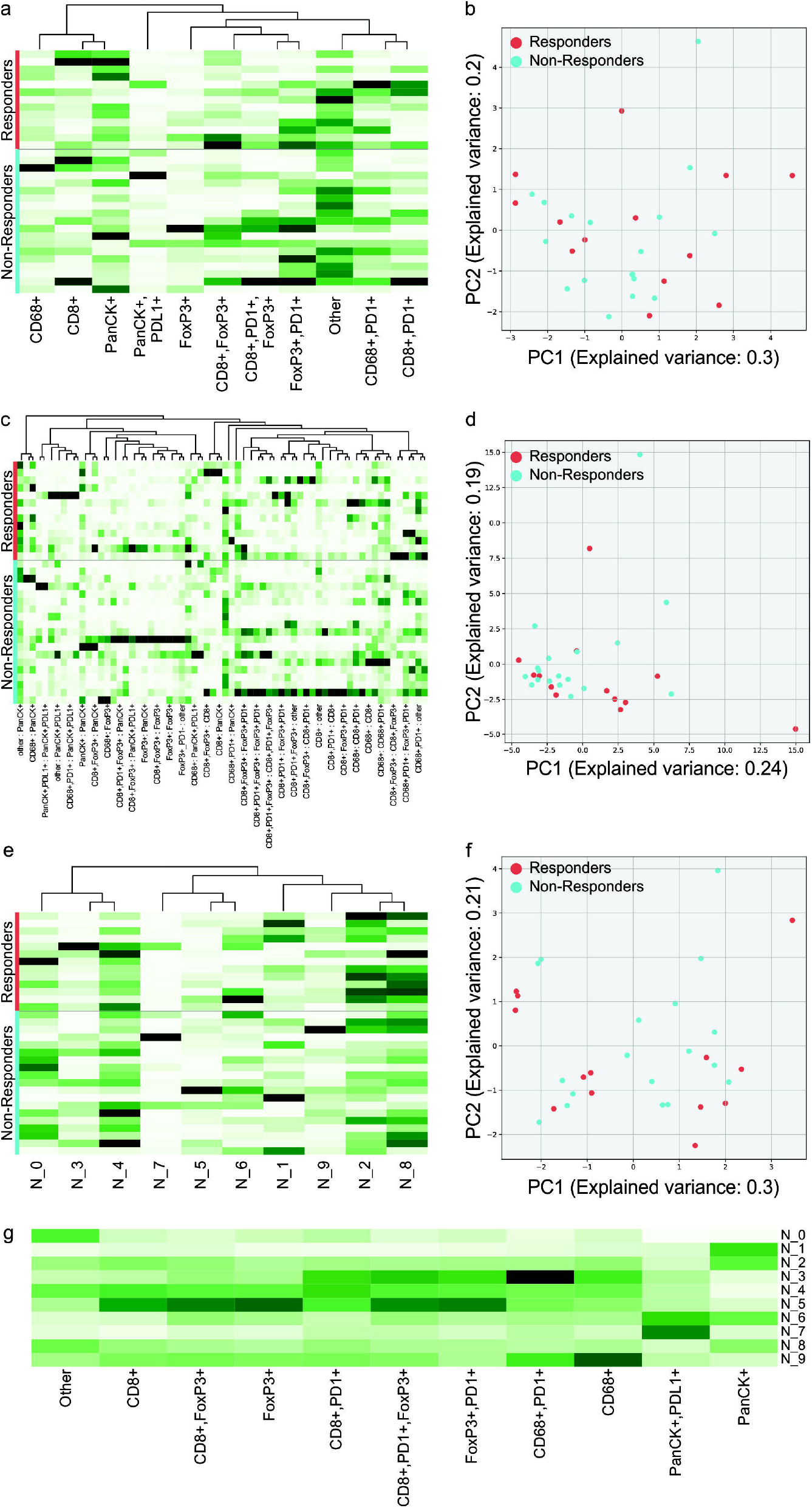
Cellular counts and spatial analysis of IO TMA by multispectral IHC. **A** Dendrogram of normalised cell frequencies of samples. **B** PCA of sample cell frequencies. **C** Dendrogram of cell-cell interaction proportions of samples (32/66 interactions labelled for legibility). **D** PCA of sample cell-cell interactions. **E** Dendrogram of neighbourhood associations in samples. **F** PCA of neighbourhoods in samples. **G** Heatmap showing relative abundance of cell interactions in each neighbourhood.

**Supp Figure 2.**
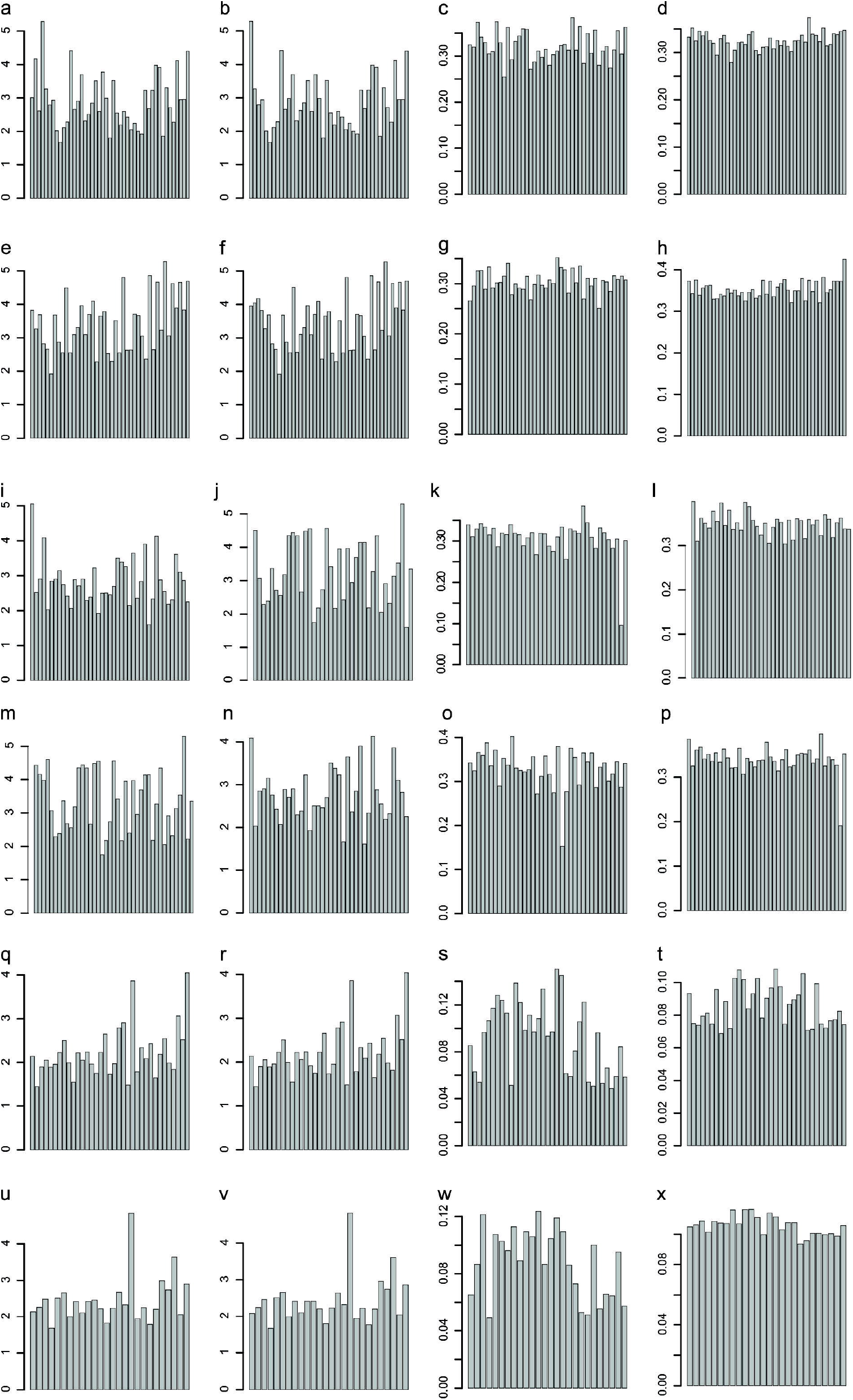
Assessment of normalisation approaches for DSP data by coefficient of variation (CV). **A, E, I, M, Q, U** Raw data (**QC**). Normalisation by: **B, F, J, N, R, V** Geometric mean of 32 internal housekeepers (**HK**). **C, G, K, O, S, W** Log2 transformation of HK data (**Log2(HK)**). **D, H, L, P, T, X** RUVg-III (**RUVg**). Each dataset was assessed independently: **A-D** SOC protein tumour. **E-H** SOC protein stroma. **I-L** IO protein tumour. **M-P** IO protein stroma. **Q-T** IO mRNA tumour. **U-X** IO mRNA tumour.

**Supp Figure 3.**
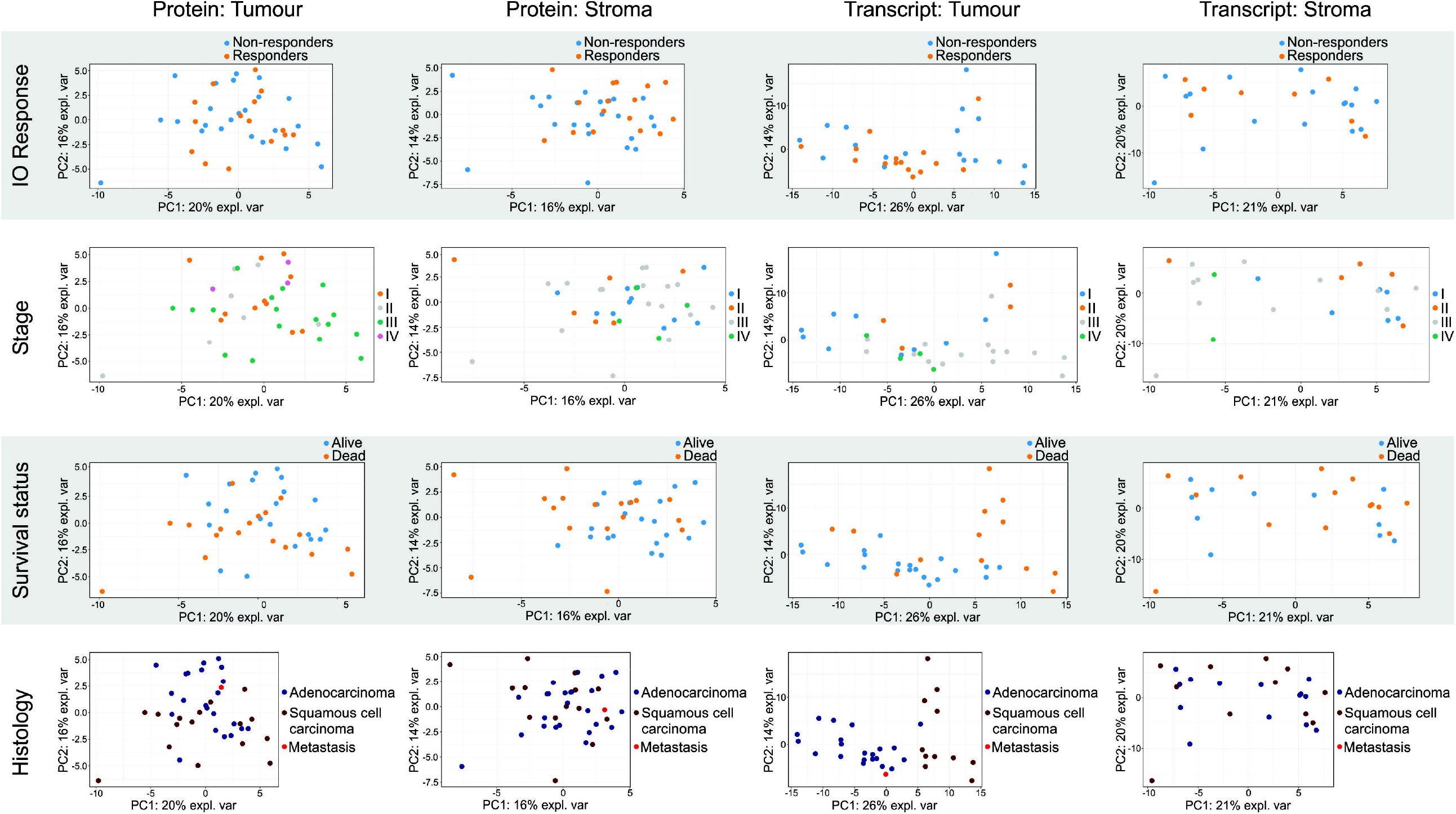
Principal component analysis (PCA) of each normalised dataset. PCA was performed for each dataset to evaluate sample association with ICI response, Stage, Survival status, and NSCLC histology.

**Supp Figure 4.**
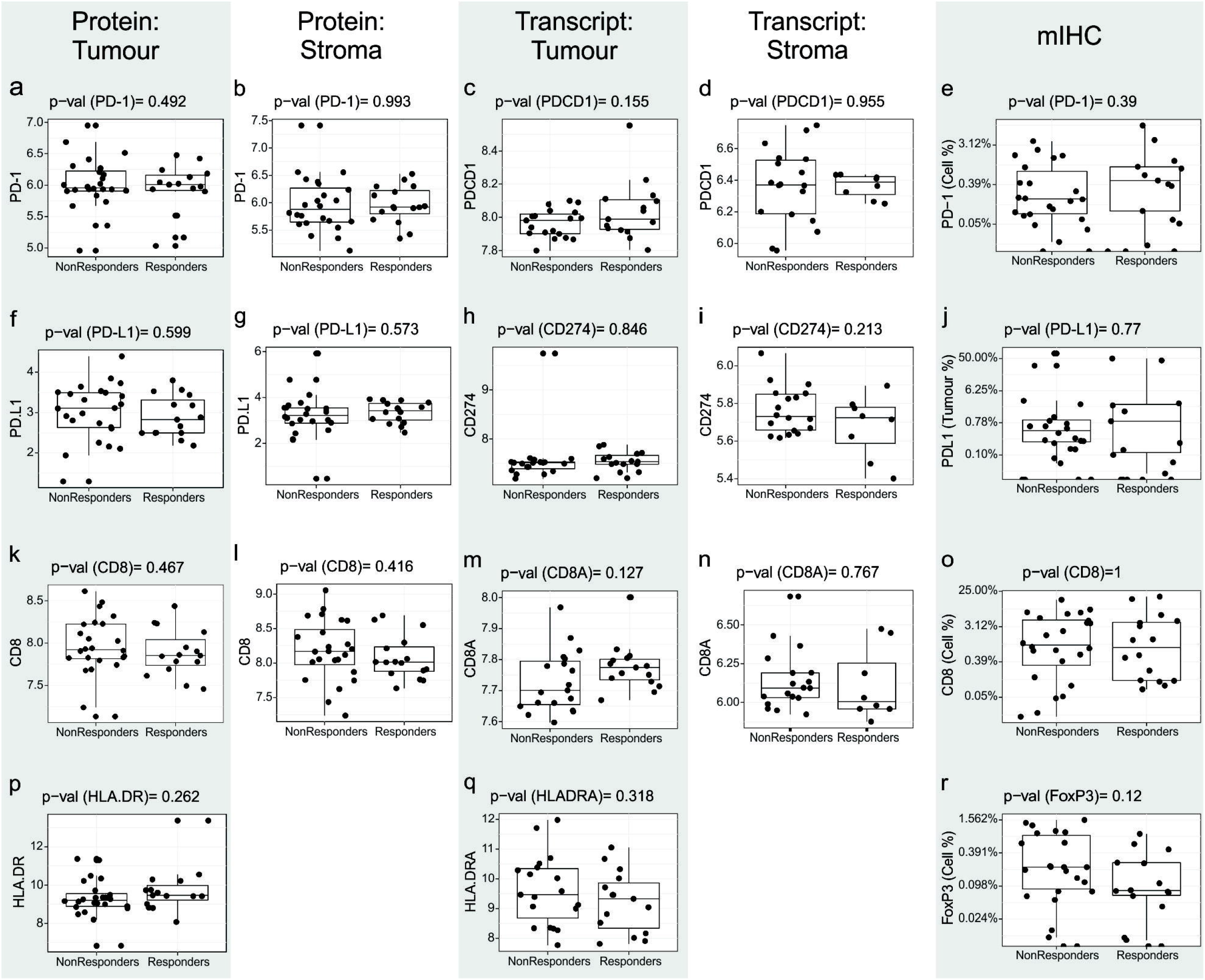
Evaluation of expression of postulated IO biomarkers in DSP and mIHC data. **A, F, K, P** IO protein tumour data. **B, G, L** IO protein stroma data. **C, H, M, Q** IO mRNA tumour data. **D, I, N** IO mRNA stroma data. **E, J, O, R** IO mIHC data. **A-E** PD-1 expression. **F-J** PD-L1 expression. **K-O** CD8 expression. **P-Q** HLA-DR in tumour only. **R** FoxP3 by mIHC only.

**Supp Figure 5.**
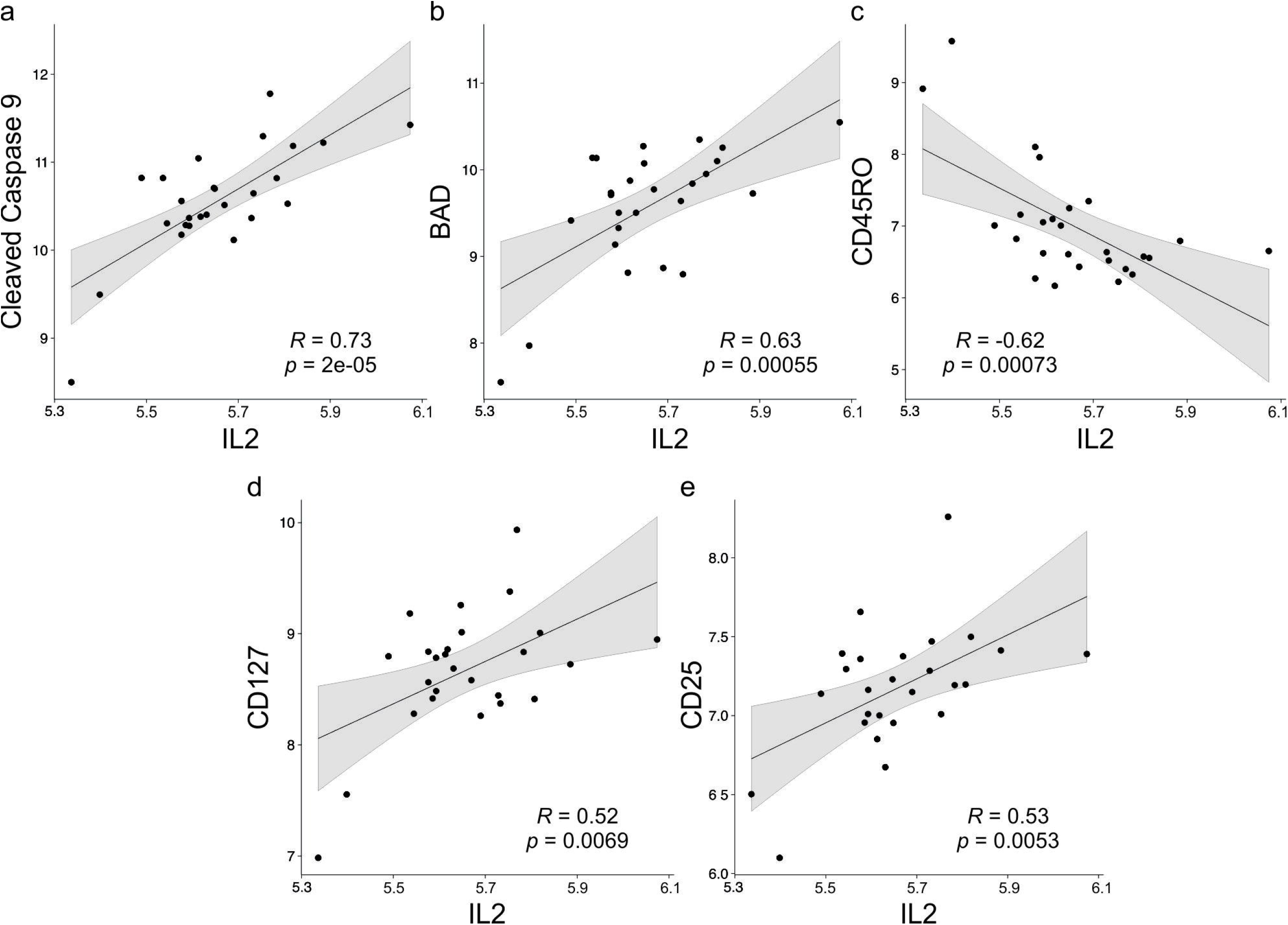
Stromal protein markers that correlate with *IL2* mRNA expression. **A** *IL2* vs Cleaved Caspase 9. **B** *IL2* vs BAD. **C** *IL2* vs CD45RO. **D** *IL2* vs CD127. **E** *IL2* vs CD25

**Supp Figure 6.**
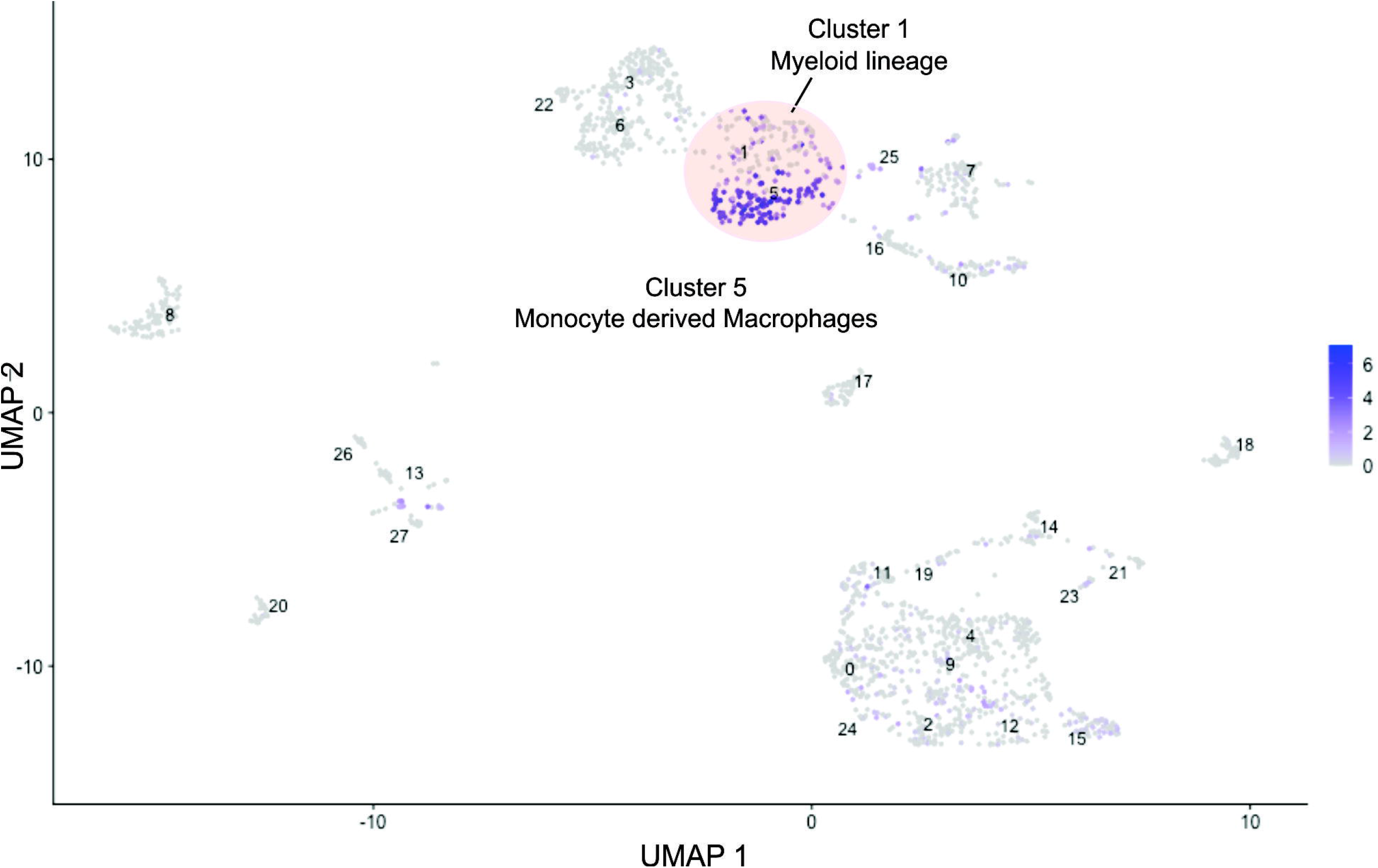
Analysis of SPP1 transcript in scRNAseq data (GSE131907) indicates enrichment of transcript from myeloid cells and monocyte derived

## Supplementary Tables

**Supplementary Table 1. Differentially expressed features of ICI response**. Differential expression was performed for protein and mRNA assays for tumour and stroma compartments.

**Supplementary Table 2. Cox Proportional Hazards survival analysis of protein and mRNA features**. Overall survival was assessed as a function of feature expression in tumour or stroma. Table indicates most highly significant survival features and their association with differential analysis.

**Supplementary Table 3. Differentially expressed gene enrichment within Nanostring Cancer Transcriptome Atlas pathway annotations**. DE genes were assessed for their representation within each pathway annotation. Table shows number of genes in each pathway, number and name of genes represented in our data.

